# Transcriptome profiling of cerebrospinal fluid in Alzheimer’s Disease reveals molecular dysregulations associated with disease

**DOI:** 10.1101/2023.11.21.23298852

**Authors:** Rhys E. De Sota, Samantha J. Khoury, Jiali Zhuang, Robert A. Rissman, James B. Brewer, Stephen R. Quake, John J. Sninsky, Shusuke Toden

**Author notes:** Contact Information: Shusuke Toden: Superfluid Dx., 259 East Grand Avenue, South San Francisco, CA 94080. Financial Support: Alzheimer’s Drug Discovery Foundation Diagnostic Accelerator: Peripheral Biomarkers Program.

## Abstract

Despite the increasing prevalence of neurodegenerative diseases, the molecular characterization of the brain remains challenging due to limited access to the tissue. Cerebrospinal fluid (CSF) contains a significant proportion of molecular contents originating from the brain, and characterizing these molecules has served as a surrogate to evaluate molecular dysregulation in the brain. Here we performed cell-free messenger RNA (cf-mRNA) RNA-sequencing on 52 human CSF samples, and further compared their transcriptomic profiles to matched plasma samples. In addition, we evaluated the molecular dysregulation of cf-mRNA in CSF between individuals with Alzheimer’s disease (AD) and non-cognitively impaired (NCI) controls. The molecular content of CSF cf-mRNA was distinct from plasma cf-mRNA, with a substantially higher number of brain-associated genes identified in CSF. We identified a large set of dysregulated gene transcripts in the CSF cf-mRNA population of individuals with AD, and these gene transcripts were used to establish a diagnostic classifier to discriminate AD from NCI subjects. Notably, the gene transcripts were enriched in biological processes closely associated with AD, such as brain development and synaptic signaling. We also discovered a subset of gene transcripts within AD subjects that exhibit a strong correlation between CSF and plasma cf-mRNA. This study not only reveals the novel cf-mRNA content of CSF but also highlights the potential of CSF cf-mRNA profiling as a tool to garner pathophysiological insights into AD.

## INTRODUCTION

Alzheimer’s disease (AD) is the most prevalent form of dementia globally, whose cytopathology is marked by dysfunction across a wide range of biological systems, including synaptic transmission, inflammation, oxidation, protein folding, and mitochondrial metabolism (1–3). While early diagnosis of AD relied exclusively on clinical indications of disease particularly memory loss, parallel biological constructs of disease including the biomarkers of the amyloid cascade hypothesis have more recently been employed. Aligned with this hypothesis, PET imaging of amyloid beta (Aβ) and tau serve as the reference for detection of disease. Established biofluid biomarkers for AD primarily center around the detection of different forms and ratios of extracellular Aβ and the intracellular formation of differentially phosphorylated tau forming neurofibrillary tangles in the brain. Biofluid Aβ and tau are measured mainly in cerebrospinal fluid (CSF) with more recent efforts detecting various forms of these amyloid cascade biomarkers in blood(4).

One of the primary limitations of conventional AD protein biomarkers is they only reflect a fraction of the complex biology involved in the disease. Indeed, there is a growing body of evidence suggesting the majority of elderly individuals with dementia often exhibit more than one hallmark pathology associated with cognitive decline (5). Individuals with AD often have concomitant cerebrovascular disease pathologies, as well as other neurodegenerative disease pathologies (6). Furthermore, genetic and molecular factors have been proposed to play a role in individuals who display AD pathologies without exhibiting dementia symptoms, a phenomenon referred to as "AD resilience" (7). Therefore, the conventional diagnostic framework may not be sufficient to fully encompass the overlapping pathologies and potential endotypes observed in AD.

Recent technological advances have enabled comprehensive molecular profiling of various diseases. Accordingly, several gene-expression profiling studies using post-mortem human brain tissue samples, have demonstrated the transcriptional profile of patients with AD differs substantially from that of non-cognitively impaired individuals (8–10). However, conducting transcriptomic profiling of brain tissue presents challenges due to the time-dependent degradation of tissue and dysregulation of gene expression at death (11, 12). Furthermore, establishing a time-controlled collection procedure for postmortem brain tissues poses significant challenges. Liquid biopsy has emerged as a promising minimal invasive alternative to surgical biopsies, especially for organs that are difficult to access (13). The advancements in next-generation sequencing (NGS) have facilitated comprehensive hypothesis-independent quantification of different types of circulating nucleic acids (14). We have developed an RNA-Seq based platform that allows accurate quantification of circulating cell-free messenger RNA (cf-mRNA) (15–19). Surveying the mRNA in the cell free compartment provides more direct insight into the involved underlying pathological processes.

By employing unbiased transcriptome profiling, we resolved the tissue and cell type of origin for circulating mRNA transcripts, utilizing well-established human tissue and single cell RNA sequencing databases (18, 20, 21). In our previous work, we conducted cf-mRNA transcriptome profiling of plasma samples from individuals with and without AD. This study identified genes dysregulated in AD compared to the non-cognitively impaired controls and demonstrated that these genes could be used to develop an AD diagnostic classifier (15). Although plasma-based analytes like cf-mRNA offer an accessible means for developing diagnostic assays and permit sampling of disease pathology reflected in circulating cells (22), they have limitations in providing molecular content from the brain. This limitation arises due to the elevated presence of circulating transcripts derived from blood cells and other organs, hindering the specificity of brain-related molecular analysis (15, 16). In contrast, CSF contains a significant portion of molecular contents originating from the brain (23). Here, we evaluated the cf-mRNA molecular dysregulation between AD subjects and non-cognitively impaired controls in 52 CSF samples and subsequently compared their transcriptomic profiles to plasma samples.

## MATERIALS AND METHODS

### Clinical specimens

We examined a total of 52 CSF specimens, comprised of 40 AD and 12 non-cognitive impaired (NCI) subjects. Specimens were collected from the University of California, San Diego and BioIVT. Further subject demographics and clinicopathological details can be found in Supplementary Table 1. Plasma sequencing data have been previously published and are accessible through the Sequence Read Archive (SRA: PRJNA574438) (15). All participants provided written informed consent, and the study approved by the institutional review boards of participating institutions.

### RNA extraction, library preparation and whole-transcriptome RNA-seq

RNA was extracted from up to 1 mL of both CSF and plasma using the QIAamp Circulating Nucleic Acid Kit (Qiagen, Hilden, Germany) and then eluted in a 15 µl volume. To serve as an exogenous spike-in control, the ERCC RNA Spike-In Mix (Thermo Fisher Scientific, Waltham, MA, Cat. # 4456740) was added to the RNA following the manufacturer’s instructions. The integrity of the extracted RNA was evaluated using the Agilent RNA 6000 Pico chip (Agilent Technologies, Santa Clara, CA, Cat. # 5067-1513), and the RNA samples were converted into sequencing libraries as previously described (16). The NGS library preparation process underwent both qualitative and quantitative analysis utilizing chip-based electrophoresis (Agilent DNA high sensitivity kit, Agilent Technologies, Santa Clara, CA), and a qPCR-based quantification kit (Roche, Basel, Switzerland, Cat. # KK4824) to quantify the libraries. RNA sequencing was carried out on the Illumina NextSeq500 platform (Illumina Inc, San Diego, CA) with paired-end sequencing of 75 cycles. For base-calling, the Illumina BaseSpace platform (Illumina Inc, San Diego, CA) with the FASTQ Generation application was used, while adaptor sequences were removed and low-quality bases were trimmed using cutadapt (v1.11). Reads shorter than 15 base pairs were excluded from subsequent sequencing analyses and read sequences exceeding 15 base pairs in length were matched against the human reference genome GRCh38 using STAR (v2.5.2b), specifically utilizing GENCODE v24 gene models. To eliminate duplicated reads, the samtools (v1.3.1) rmdup command was utilized. Finally, RSEM was used to calculate gene-expression levels from the de-duplicated BAM files (v1.3.0).

### Tissue and cell type-associated gene identification

The Genotype-tissue expression (GTEx) database is a comprehensive resource for tissue specific gene expression (24), and the database was used to identify organ-specific genes (i.e., tissue-associated). GTEx tissue-associated genes are defined as genes showing substantially higher expression in a particular tissue compared to other tissue types. We obtained tissue-associated transcriptome expression levels from two public databases: GTEx (https://www.gtexportal.org/home/) for gene expression across 51 human tissues (24) and Blueprint Epigenome (http://www.blueprint-epigenome.eu/) for gene expression across 56 human hematopoietic cell types. For each individual gene, we ranked the tissue by their expression of that particular gene, and when gene expression in the top tissue (cell-type) was greater than 20-fold than all the other tissues, the gene was considered to be ‘’associated’’ with the top tissue. The list of brain and liver genes is shown in Supplementary Table 2. Brain-tissue associated gene expression levels were obtained from the Human Protein Atlas database (downloaded March 15^th^, 2022) (25), and the HPA “elevated-genes” are considered to be brain-associated. To decipher and label cell-type specificity of gene transcripts, we implemented the PanglaoDB database on our list of brain-associated genes (26). The expression levels of cell type-associated genes were normalized based on their tissue specificity.

### Differential expression analysis, pathway analysis, NMF and classifier development

Differential expression analysis was performed using the Wilcoxon rank-sum test with TPM counts as input (Python 3.10.9). Specifically, a two-sided Wilcoxon rank-sum test was used in order to limit high FDR from our large sample size, and an unadjusted p-value cutoff of 0.05 was used to define differentially expressed genes (27). Gene Ontology, KEGG and Reactome Pathway analyses were conducted using Metascape (28), with *H. sapien* as the input and analysis species. A list of differentially expressed gene IDs were uploaded to Metascape in order to determine highly enriched biological pathways. Ingenuity Pathway Analysis (IPA) (Qiagen, Ver. 101138820) was performed using differentially expressed genes as input.

Transcriptome decomposition analysis was performed using non-negative matrix factorization (NMF) as described previously (15). In brief, NMF was performed to decompose normalized gene-expression profiles from cfmRNA into 5 components. In NMF decomposition, genes sharing similar expression patterns across samples are grouped together in an unsupervised manner. NMF decomposition was performed using the scikit-learn Python machine-learning library. Genes with >40% loading attributable to a particular component were considered enriched in the component. We performed subject grouping by performing hierarchical clustering on the coefficient matrix W. Hierarchical clustering was implemented using the Python library SciPy (v1.3.0) class scipy.cluster.hierarchy.linkage with parameters method = “average” and metric = “correlation.”

To develop a classifier, we used logistic regression analysis with L2 regularization within the scikit-learn Python library (29). Hyperparameters were determined with stratified 5-fold cross-validation and the final altered parameters include solver = ‘liblinear’, penalty = ‘l2’, and C = 1.0. Additionally, genes were ranked based on the absolute value of the logistic regression coefficient in the training set and genes with highest coefficient were used to derive minimal gene classifiers. For specific analyses, principal component analysis (PCA), and hierarchical clustering were performed using sklearn.decomposition.PCA and seaborn.clustermap, respectively. Additionally, for PCA and NMF, TPM data was scaled using StandardScaler and MinMaxScaler from sci-kit learn, respectively.

### Statistical analysis

Subject characteristics were summarized using mean and standard deviation or frequency and percentage, and all statistical analyses were performed using Python version 3.10.9. (30). Risk scores derived from the gene-classifier multivariate logistic regression model were used to plot receiver-operating-characteristic (ROC) curves and calculate area under the curves (AUC). The limited number of subjects with AD prevented creation of a separate validation data set. Therefore, stratified 5-fold cross-validation was used to assess potential out of sample performance variation. Assessments involving two groups were evaluated using Wilcoxon-Rank-sum test (Python scipy.stats.mannwhitneyu).

## RESULTS

### CSF cf-mRNA contains a higher proportion of brain-associated transcripts compared to plasma cf-mRNA

While we and others have successfully conducted plasma/serum cf-mRNA-sequencing in multiple diseases (15–19, 31–35) and examined the molecular content of cf-mRNA in clinical samples, the molecular profile of CSF cf-mRNA remained unclear. We conducted RNA-sequencing on 52 CSF specimens, out of which 13 had corresponding matched plasma samples that were sequenced in a procedurally equivalent manner. (Figure 1A) (15). In order to assess the overall molecular profile of CSF cf-mRNA compared to plasma cf-mRNA, we conducted principal component analysis (PCA) with CSF and plasma samples of 13 pairs of matched plasma and CSF samples. PCA analysis revealed that plasma and CSF formed distinct clusters, indicating that the composition of CSF and plasma are substantially different (Figure 1B). Additionally, unsupervised hierarchical clustering reiterated that CSF and plasma segregate based on fluid type (Figure 1C). Next, we assessed the composition of tissue-associated transcripts in CSF and plasma using the GTEx database for reference (Figure 1D). To ensure a comparable analysis, we assessed tissue-associated genes of both the brain and liver in subjects with matched CSF and plasma samples (n = 13) (Supplementary Figure 1A). In the CSF, we detected the highest number of transcripts originating from the brain, followed by the skin and liver, while the highest number of transcripts in plasma originated from the liver, followed by the brain and muscle.

**Figure 1:**
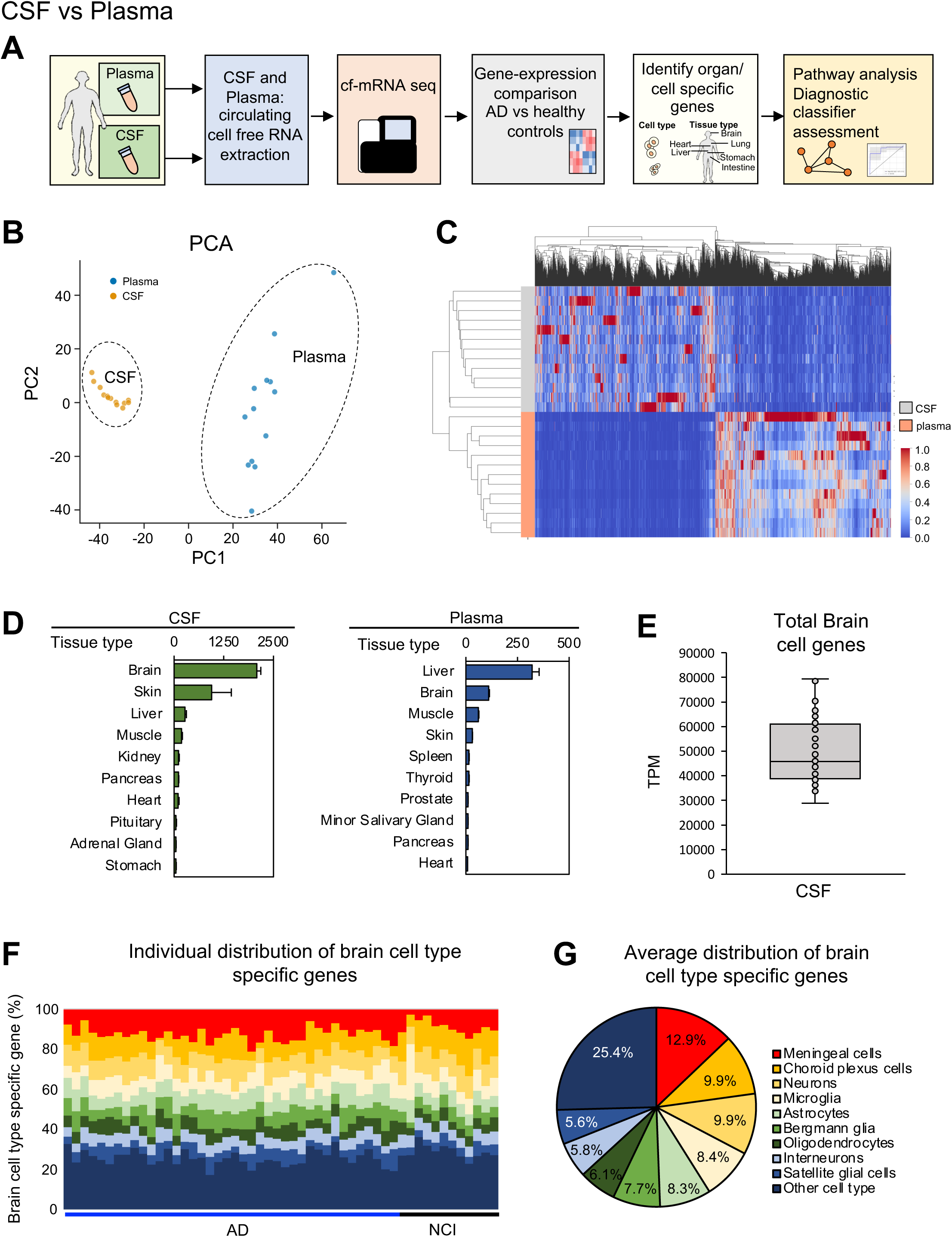
CSF and plasma cf-mRNA exhibit distinct molecular compositions. A) Study design schematic. B) PCA plot of all samples used in the study. Plasma and CSF populations are identified using dotted circle. C) Heat map of differentially expressed genes (NCI vs AD). D) Ranked quantification of tissue associated transcripts in CSF (left) and plasma (right) (All samples). E) Average number of brain cell genes detected in CSF. Average total brain cell genes in CSF. F) The distribution of genes associated to brain cell types for CSF samples from individuals with AD and those without cognitive impairment (NCI). G) Average distribution of genes associated to brain cell types for CSF.

Next, we utilized the Panglao Database to determine the overall count of brain cell genes present in CSF (Figure 1E). The average CSF sample contained more than 40,000 transcripts per million (TPM) of brain cell associated genes. Following this, we assessed the proportion of genes associated with brain cell types as a percentage and analyzed the makeup of brain cell-type associated genes within the CSF (Figure 1F). Within the CSF, the most prevalent cell types identified were meningeal cells, choroid plexus cells and neurons (Figure 1G), and approximately 17% of transcripts are associated with resident immune cells (e.g. microglia and astrocytes). Our data reveals a distinct molecular composition of CSF with notable enrichment of brain-associated genes.

### Identification of brain-associated differentially expressed genes

To examine the cf-mRNA transcriptomic profile of subjects with AD in comparison to NCI, we performed differential expression analysis on 40 AD samples and 12 NCI samples. Hierarchical clustering analysis revealed a majority of AD subjects formed into two distinct clusters and, similarly, NCI individuals were grouped into two clusters (Figure 2A). The differential expression analysis identified 1943 differentially expressed genes. Of which 955 genes were upregulated and 988 genes were downregulated (Figure 2B) (the term “upregulated” and “downregulated” are used to describe changes in the number of RNA molecules in the circulation of subjects with AD compared to NCI). We then evaluated the individual genes that are most highly dysregulated (Wilcoxon rank-sum test) (Figure 2C, Supplementary Figure 2A). Specifically, by utilizing the Human Protein Atlas, we identified that three of the top ten upregulated genes, namely CKB, CDH7, and TMEM63C, exhibited predominant expression in the brain. Next, we utilized the differentially expressed genes as input to conduct pathway analyses. The IPA analysis showed that the CREB signaling in neurons as the most dysregulated pathways for the AD upregulated genes, while SRP dependent co-translational protein targeting to membrane was the most significant pathway for AD downregulated genes (Figure 2D). Physiological system development and function subcategory of IPA indicated that “nervous system development and function” was the most significant pathway for both up and downregulated genes (Supplementary Figure 2B). These analyses indicated that cf-mRNA genes that are differentially expressed in AD were brain-associated. Similarly, the Gene Ontology analysis revealed that among the most significant pathways for upregulated genes, several were related to synaptic signaling (Supplementary Figure 2C). On the other hand, for downregulated genes, the most prominent pathway was associated with cytoplasmic translation of mRNA (Supplementary Figure 2B). In line with the Gene Ontology analysis, both KEGG and Reactome pathway analyses also highlighted "neuroactive ligand-receptor interaction" and "neuronal system" as among their top enriched pathways (Supplementary Figure 2E-D). This suggests the upregulated genes in AD are linked to neuronal dysregulation, further reinforcing the connection between these genes and the neurological alterations observed in the disease. Furthermore, through DisGeNET enrichment analysis (36), it was determined that the upregulated genes in AD exhibited associations with various other neurodegenerative diseases (Figure 2E). We also discovered that 10.8% of all dysregulated genes in AD subject samples are genes enriched in the brain, as referenced from the Human Protein Atlas (Figure 2F).

**Figure 2:**
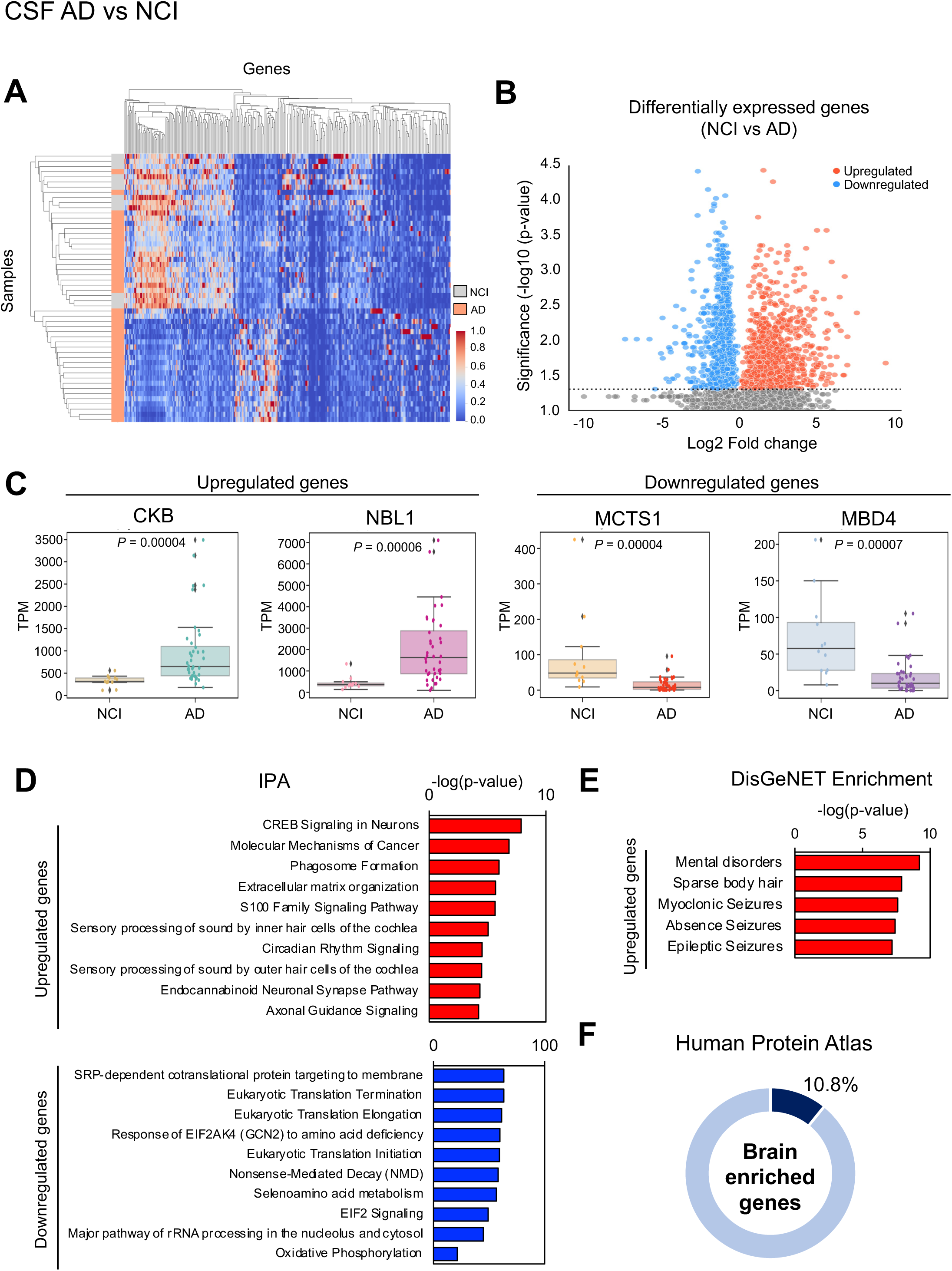
Identification of AD-associated differentially expressed CSF cf-mRNA genes. A) Heat map of differentially present gene transcripts (NCI vs AD). Subject types are shown on y-axis. B) A volcano plot showing the dysregulation of cf-mRNAs in subjects with AD compared to that of NCI (Wilcoxon rank-sum test). C) Examples of the most upregulated (left) and downregulated (right) genes. D) Top IPA canonical pathways for genes that are dysregulated in AD, upregulated and downregulated genes are used as input. E) Top DisGeNET enrichment pathways for AD upregulated genes. F) Percentage of brain enriched genes (Human Protein Atlas) in differentially present genes (AD vs NCI).

### The utilization of cf-mRNA profiling to identify potential AD subtypes

To further interrogate the CSF cf-mRNA transcriptome of AD subjects, we evaluated whether these differentially expressed genes are organized into functionally related clusters. We performed non-negative matrix factorization (NMF) on the differentially expressed genes which resulted in 5 clusters (205, 255, 461, 177 and 236 genes respectively) (Figure 3A). Next, we used genes associated with each cluster to perform multiple gene enrichment analyses (Figure 3B and Supplementary Figure 3A-D). The IPA analysis showed that cluster 1, 2 and 3 had genes that are associated with “nervous system development and functions” (Figure 3B). Cluster 3 appears to be associated with dysregulation in neuronal/synaptic system (Supplementary Figure 3C and D), while cluster 4 is associated with DNA-damage/cell cycle responses (Supplementary Figure 3B). Moreover, when compared to the "brain elevated genes" reference from the Human Protein Atlas, Clusters 2 and 3 exhibited a significantly higher proportion of brain-elevated genes, accounting for 16.7% and 16.8% of their respective gene sets (Figure 3C). Next, we assessed whether there might be distinct AD endotypes based on the differentially expressed genes by applying NMF to the subjects (Figure 3D). We identified 5 different patient groups, where group 1, 2 and 3 corresponded to Cluster 1, 2 and 3 from the differentially expressed gene based NMF (Figure 3A). In particular, the majority of the AD subjects were classified into Groups 1 and 2.

**Figure 3:**
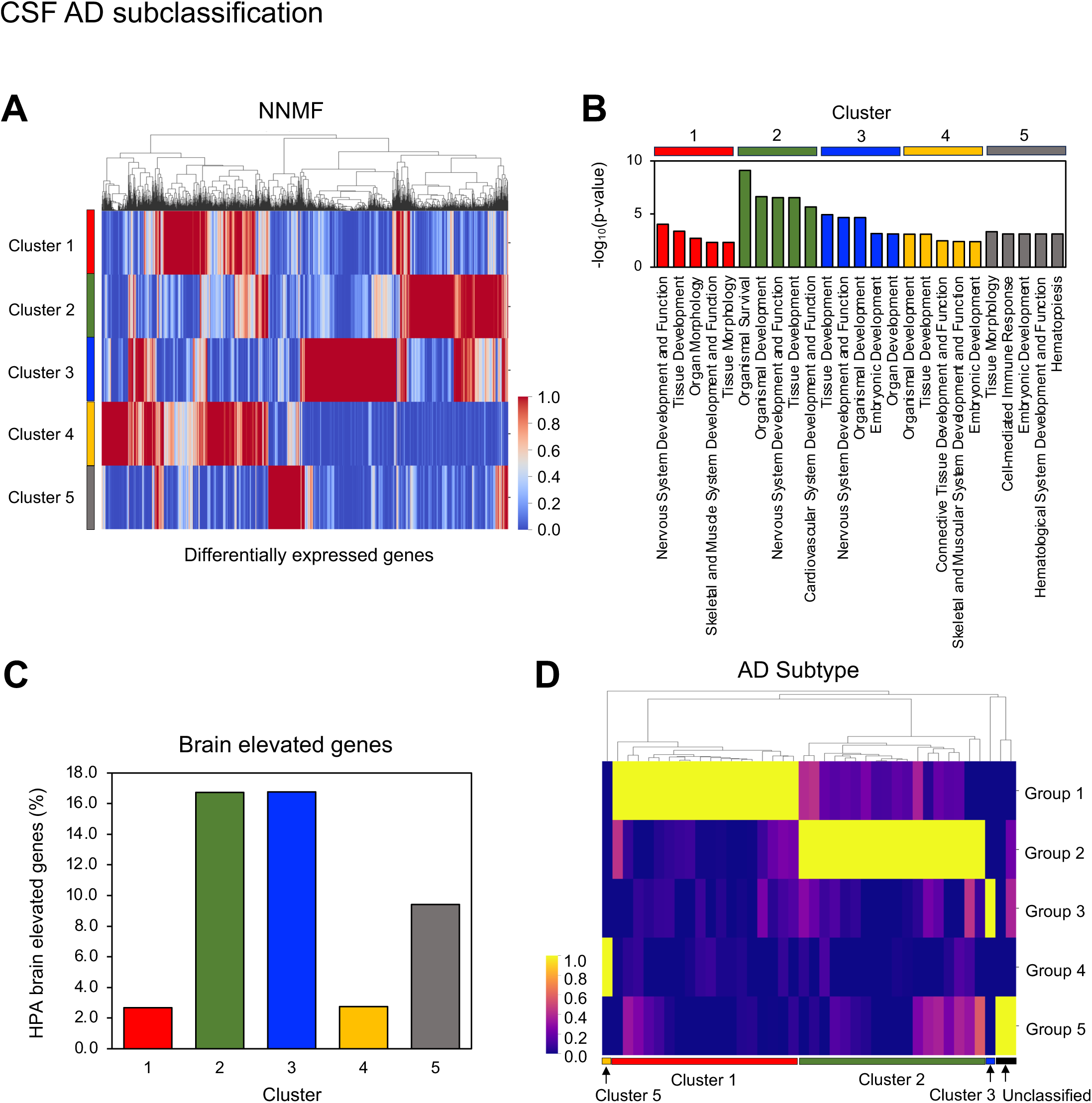
AD Molecular Clusters. A) Consensus matrix NMF clustering of genes that are differentially present in cf-mRNA of subjects with AD compared to NCI. 5 major cluster subtypes are labelled. B) IPA Physiological System Development and Function analysis of each cluster (top 5 pathways are shown). C) Number of brain elevated genes in each cluster. D) Consensus matrix NMF clustering of AD subjects using genes that are differentially expressed in cf-mRNA of subjects with AD compared to NCI.

### Cf-mRNA biomarkers for moderate AD to mild AD differentiation and other AD associated biomarkers

Next, we explored the presence of genes exhibiting differential expression between the clinical stages of dementia. We categorized subjects into mild and moderate dementia stages based on their mini-mental state examination (MMSE) scores for cognitive impairment (AD mild: 26-19, AD moderate: 18-10) (37). Hierarchical clustering analysis revealed the majority of moderate dementia subjects showed distinctively different molecular profiles compared to mild dementia subjects (Figure 4A). We identified 906 differentially expressed genes, among them 655 were upregulated and 251 were downregulated (Figure 4B). We then evaluated the most highly dysregulated genes individually (Supplementary Figure 4A). We identified FBXO9 and SEZ6L2 as the genes that are most highly upregulated in moderate dementia compared to mild dementia (Wilcoxon rank-sum test). In contrast, CNR1 and GPD2 were the most downregulated genes in moderate dementia compared to mild dementia (Wilcoxon rank-sum test) (Figure 4C). Of note, both FBXO9 and SEZ6L2 have been shown to be involved in proteostasis (38, 39), a known feature of AD (40). In addition, SEZ6L2 has been shown to have implications in neuronal and especially motor function development (41, 42). Furthermore, high expression of CNR1 in the brain reduced neuronal injury in hippocampus in an AB25-35 induced rat model (43), while GPD2 has been shown to be dysregulated in a neurological disorder *in vivo* (44).

**Figure 4:**
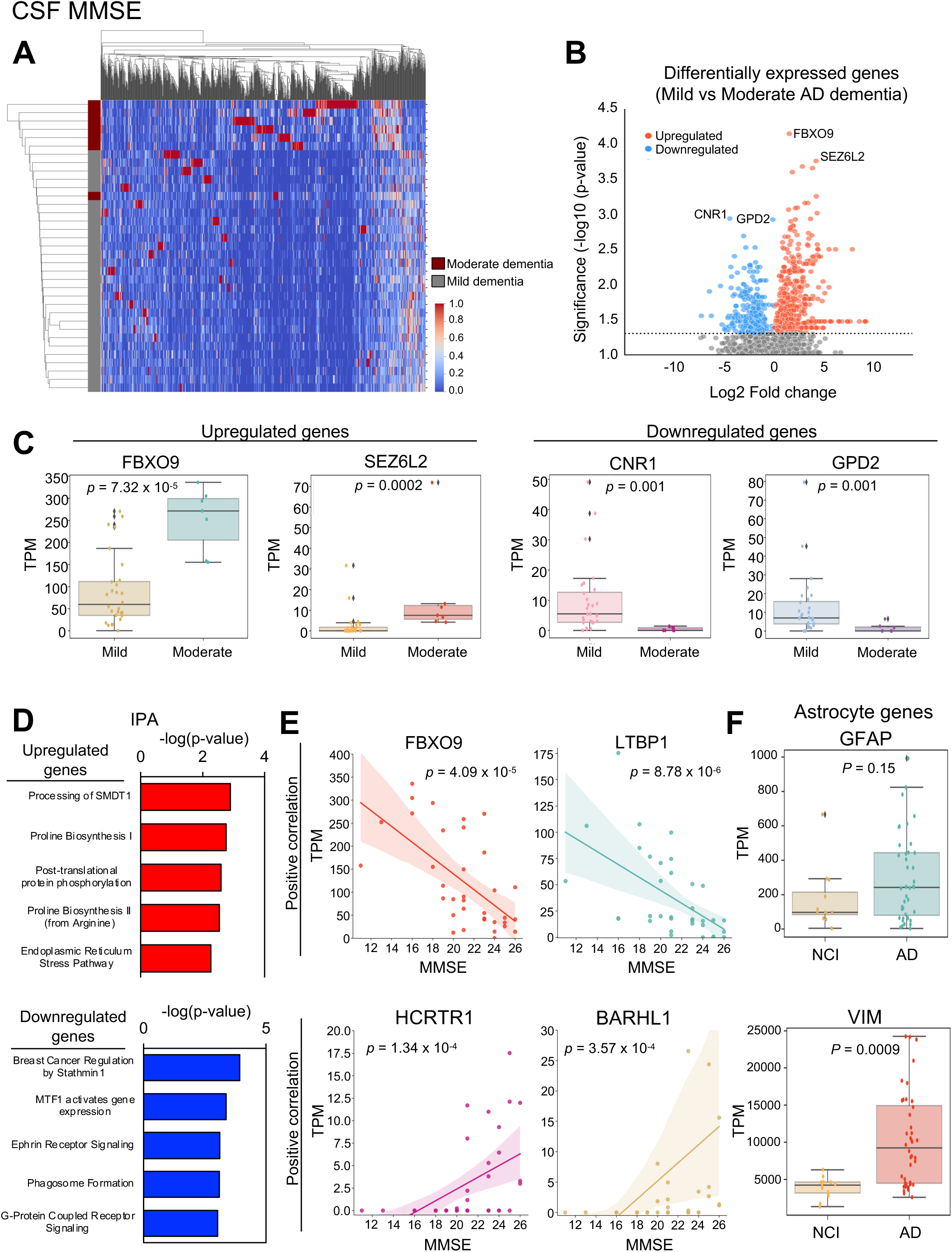
Molecular profiles of mild vs moderate dementia in AD for CSF cf-mRNA. A) Heat map of differentially present genes (Mild vs Moderate dementia). Subject types are shown on y-axis. B) A volcano plot showing the dysregulation of cf-mRNAs in subjects with mild dementia compared to that of moderate dementia (Wilcoxon rank-sum test). C) Examples of the most upregulated (left) and downregulated (right) genes. D) Top IPA canonical pathways for genes that are dysregulated in AD, upregulated and downregulated genes are used as input. E) Top correlated genes with MMSE scores (positive correlation (top), negative correlation (bottom). F) The expression levels of Astrocyte activity associated genes.

Next, we performed pathway analysis using genes that are dysregulated in moderate AD compared to mild AD. IPA canonical pathway analysis revealed that top pathways of genes upregulated in moderate dementia compared to mild dementia was processing of SMDT1 (Figure 4D) and appear to have more genes that are associated with “nervous system development and functions” (Supplementary Figure 4C). Gene Ontology analysis revealed that several top pathways of genes upregulated in moderate dementia compared to mild dementia were cilium associated pathways (Supplementary Figure 4D). Prior studies have indicated a potential link between AD and ciliopathies (45, 46). This insight implies that pathways associated with cilia could play a crucial role in the progression of AD. Given our findings suggesting molecular disparities in CSF cf-mRNA between mild and moderate dementia, we extended our investigation to explore correlation between CSF cf-mRNA genes and the MMSE score. We ranked genes based on both nominal p-value and correlation coefficient (Rho) to identify genes with high correlation between CSF and plasma (Supplementary Figure 4B). Interestingly, the gene with the highest negative correlation between MMSE score and CSF expression was FBXO9, followed by LTBP1 (Figure 4E). In contrast, HCRTR1 and BARHL1 were the genes exhibiting the strongest positive correlation between their respective CSF cf-mRNA expression and the MMSE score (Figure 4E). Collectively, our data suggest that not only can cf-mRNA profiling identify genes associated with AD diagnosis but can identify genes that are associated with cognitive impairment within an AD subject set.

In addition, we examined whether cf-mRNA gene expression dysregulation correlates with brain MRI scores. We identified MRI parameters that differed between mild to moderate dementia (Supplementary Figure 5A). Subsequently, we identified cf-mRNA genes that correlate highly with these MRI parameters (genes with highest Rho value shown, positive correlation Supplementary Figure 5B, negative correlation Supplementary Figure 5C). Although a larger data set is required to evaluate whether cf-mRNA genes can recapitulate MRI changes in the brain, there appear to be genes that reflect morphological changes in the brain.

One of the key advantages of using RNA-Seq for comprehensive AD profiling is its ability to interrogate the entire transcriptome and assess the diagnostic potential of previously reported AD-associated genes. Reactive astrocytes have been associated with AD diagnosis and prognosis (47, 48). Specifically, GFAP is the most commonly utilized marker for identifying reactive astrocytes (49). We quantified the cf-mRNA levels of GFAP, as well as VIM, which is another recognized marker for reactive astrocytes (50). Our results showed upregulation of VIM in CSF of AD subjects compared to NCI (Figure 4F). Moreover, we observed a trend of elevated GFAP expression levels in the CSF of moderate dementia compared to mild dementia, however, there were no significant differences between the groups in terms of VIM expression between moderate and mild dementia (Supplementary Figure 6A). We found that there was a trend of correlation between MMSE and GFAP expression levels, though (Supplementary Figure 6B). Collectively, our data demonstrates that cf-mRNA profiling enables simultaneous quantification of multiple genes and biological pathways.

### Cf-mRNA disease classifiers for AD

Considering that we have identified a group of genes that are differentially expressed in AD, we examined whether we could use these differentially expressed genes to establish a disease classifier. We used a total of 52 CSF samples, consisting of 40 from AD subjects and 12 from NCI individuals (Figure 5A). Our analysis involved the identification of differentially expressed genes between AD and NCI groups, which were then utilized as input data. Subsequently, we generated multiple area under the curve (AUC) values, using logistic regression analysis, to distinguish AD from NCI, utilizing different numbers of input genes by eliminating genes with the lowest coefficients (Figure 5B). When assessing the performance of these classifiers, we found that a combination of 10 genes exhibited the highest efficiency in discriminating AD from NCI subjects, with AUC value of 0.97. (Figure 5C, Supplementary Figure 7A). Furthermore, all 10 genes had relatively high TPM counts and 8 out of 10 genes were downregulated in AD (Figure 5D). In addition, we evaluated whether we could develop a robust diagnostic classifier to distinguish mild AD and moderate AD subjects. We initially assessed the robustness of the classifier by utilizing a different number of input genes, then eliminating genes with the lowest coefficients (Figure 5E). We found that a set of 50 genes exhibited the highest efficiency in distinguishing between mild dementia and moderate dementia while utilizing the fewest number of genes (AUC = 1.00) (Figure 5F, Supplementary Figure 7B and C). While the sample size of our study is small, and further validation of these classifiers in an independent validation cohort is necessary, our exploratory data suggest that CSF cf-mRNA holds promise as a valuable resource for the development of cf-mRNA-based biomarkers for AD.

**Figure 5:**
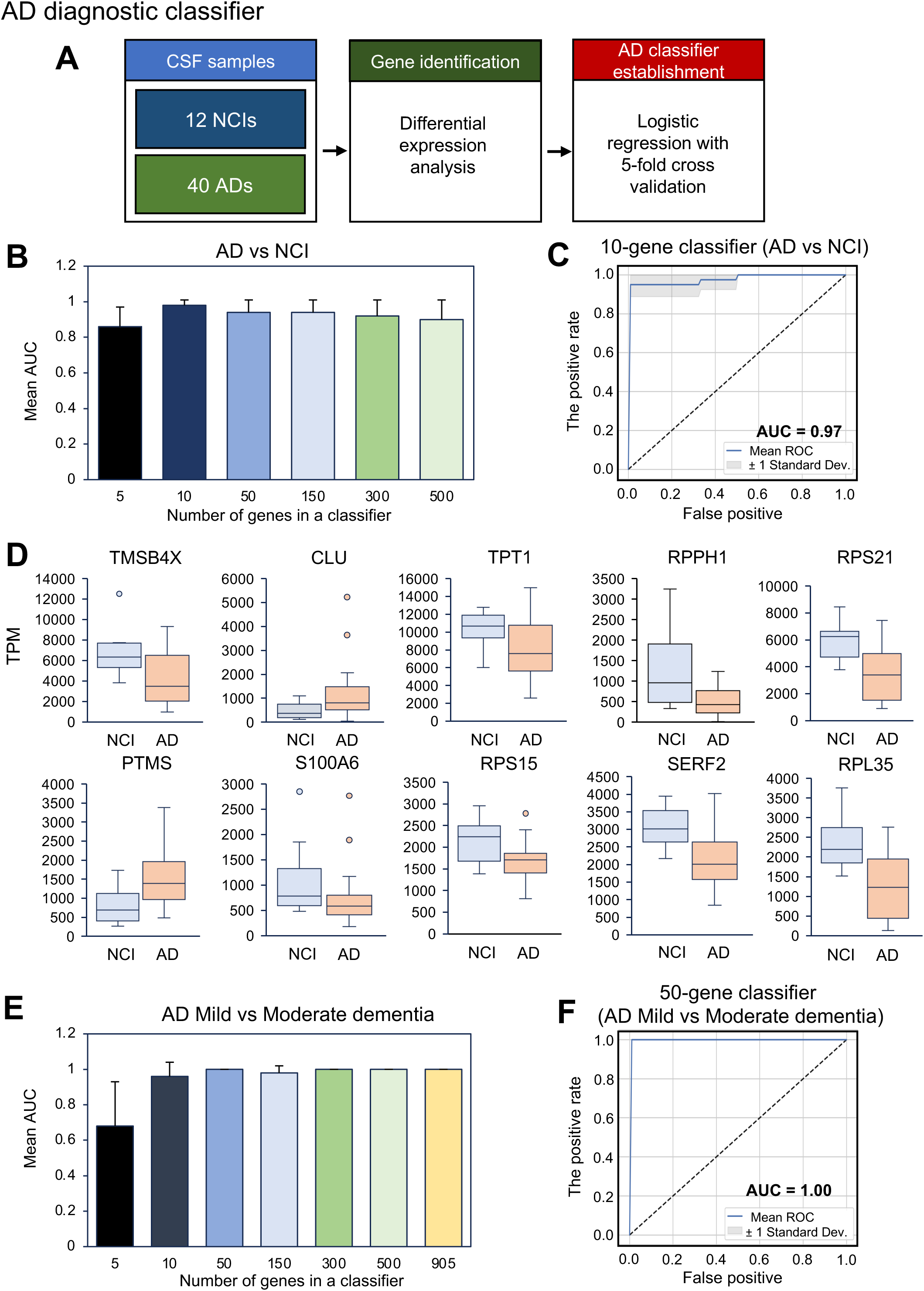
Establishing AD disease classifiers using cf-mRNA genes. A) Schematics of the analysis. B) The average AUC for genes included in the AD disease classifier (AD vs NCI). Genes with highest coefficients remained in the classifier. C) ROC curve of 10-gene cf-mRNA AD classifier that discriminates from NCI. D) The expression levels of individual genes used in the 10-gene AD classifier. E) The average AUC for genes included in the moderate dementia diagnostic classifier (moderate vs mild dementia). Genes with highest coefficients remained in the classifier. F) ROC curve of 50-gene cf-mRNA moderate dementia classifiers that discriminates from mild dementia.

### Identification of plasma cf-mRNA genes that correlate with CSF cf-mRNA

While investigating molecular dysregulation of the brain through CSF analysis is a valuable approach, it is important to recognize that CSF is not an optimal source for developing a widely accessible biomarker due to the challenges related to invasive collection. Moreover, the molecular content of the brain is released into the bloodstream directly through three blood brain barriers (blood-brain, blood CSF and the arachnoid space) (23) (Figure 6A). Hence, we proceeded to investigate whether there are plasma cf-mRNA genes that demonstrate a strong correlation with CSF cf-mRNA. Initially, we evaluated the extent of overlap among differentially expressed genes between CSF and plasma cf-mRNA when comparing individuals with AD and NCI (Figure 6B). Using all the differentially expressed genes we identified in our previous study (15), we identified 300 genes that overlapped between CSF and plasma (equivalent to 13.6% of CSF differentially expressed genes). Next, we used 13 matched plasma and CSF samples to perform Spearman’s correlation analysis assessing which plasma genes have the highest correlation to CSF (Figure 6C). We compared genes that showed high correlation between CSF and plasma (rho > 0.6 and < −06) with those that exhibited differential expression between AD and NCI in the CSF (Figure 6D). Among the genes that demonstrated a positive correlation between CSF and plasma and that are differentially expressed in AD, RGS12 and CIR1 were the two most highly correlated. On the other hand, MEGF8 and ZC4H2 emerged as the most highly negatively correlated genes while differentially expressed between CSF and plasma (Figure 6E). These data highlight that there are a group of plasma cf-mRNA genes that may correlate with CSF and may potentially inform and be used as surrogates of CSF cf-mRNA biomarkers.

**Figure 6:**
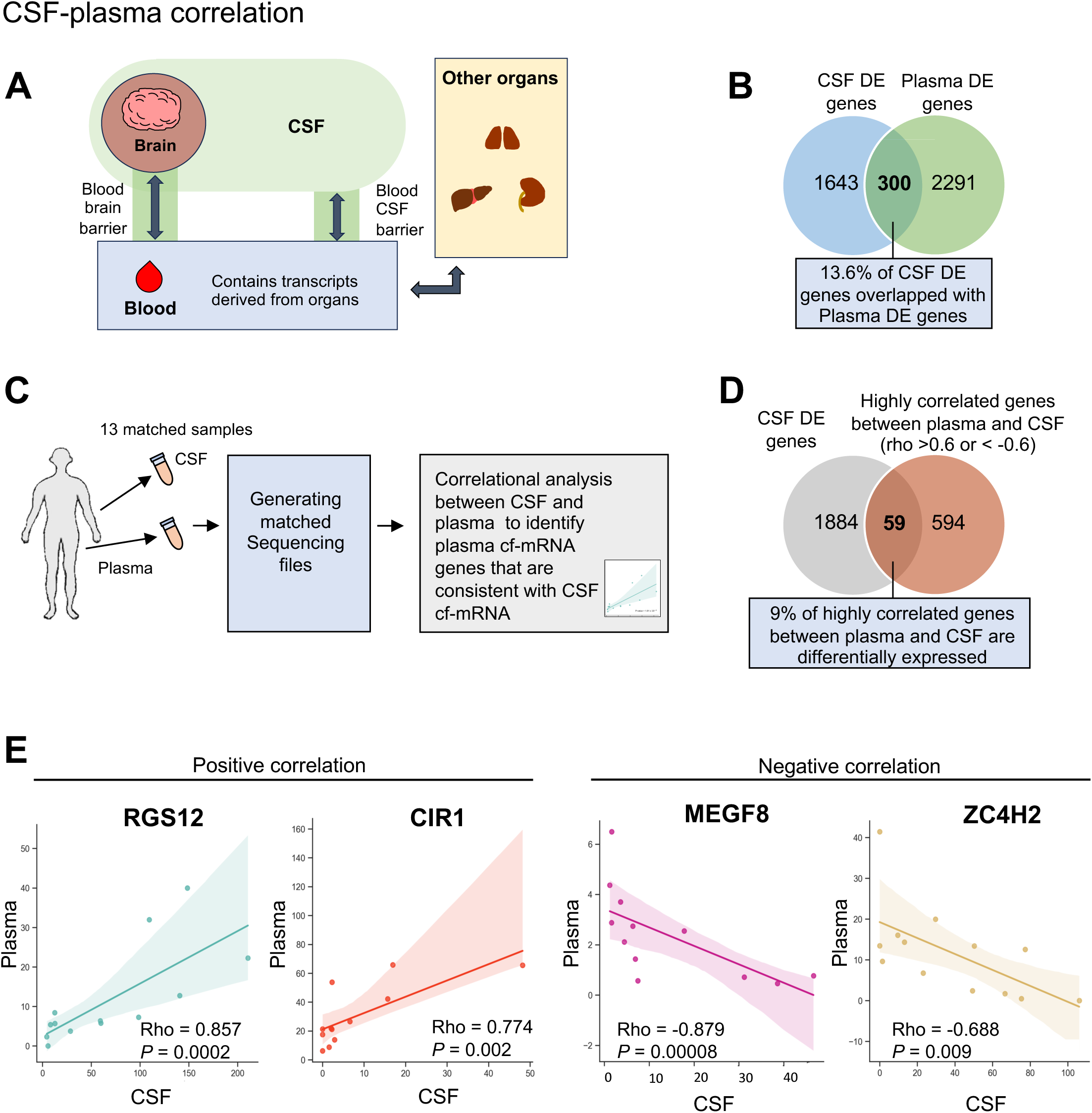
Plasma cf-mRNA genes that correlates with CSF cf-mRNA genes. A) Schematic of how transcripts from the brain are released to CSF and plasma. B) Overlap of genes that are differentially present between CSF and plasma. C) Schematic of the analysis. D) Overlap of genes that are differentially expressed present AD and NCI in CSF cf-mRNA and CSF genes that are highly correlated with plasma (rho > 0.6 or <-0.6). E) Top positively correlated genes between CSF and plasma that are differentially expressed between AD and NCI (left) Top negatively correlated genes between CSF and plasma that are differentially expressed between AD and NCI (right).

## DISCUSSION

Despite being widely recognized as a significant and increasing consequence of aging, AD continues to present major challenges, primarily due to the limited understanding of the molecular mechanisms compared to other common complex diseases (51). We were motivated to carry out this cf-mRNA study with matched CSF and plasma for three primary reasons. First, we wanted to investigate the molecular profile of the CSF cf-mRNA, a distinct bioactive compartment, especially given its proximity to the brain and lack of additional contributing factors from the systemic vasculature. Second, an unbiased survey of the cf-mRNA transcriptome may shed new light on the underlying mechanisms of disease which may in turn inform present and future pharmacotherapy interventions. Third, as CSF sampling for Aβ and tau tests only requires a fraction of the typical CSF sample volume, we reasoned that utilizing the remaining CSF with comprehensive molecular profiling may in the future provide an opportunity to complement the low dimensional protein perspective of these analytes. Considering that lumbar punctures to obtain CSF can be both painful and costly, extracting the maximum amount of information from each sample to guide patient management represents a valuable commitment to both the patient and the healthcare system. As such, cf-mRNA profiling may provide molecular insights beyond the conventional Aβ and tau tests in clinical samples.

In order to gain insight into the underlying mechanisms of AD, multiple molecular profiling techniques have been used to investigate the proteome as well as the transcriptome of small RNAs in CSF (52–55). For proteomic approaches, both immunoassay-based proximity extension assay (52) and mass-spectrometry (53) have been widely used to examine the molecular profiles of AD. Similarly, multiple studies have examined the microRNA profiles of AD CSF using multiplex PCR (54, 55). While these studies have offered valuable insights into the molecular dysregulations occurring in AD, it is important to note that they did not employ unbiased molecular profiling platforms such as RNA-sequencing. In our study, we utilized NGS-based cf-mRNA RNA-seq to characterize the whole transcriptome of plasma samples acquired from individuals with AD. Progress in clinical-grade NGS over last decade promises to facilitate introduction of clinical-grade RNA-Seq in the future for accurate detection of low expressing genes (56–58). To the best of our knowledge, this is the first study to conduct a comprehensive transcriptomic profiling of CSF for a neurodegenerative disease. Our findings indicate that CSF comprises notably elevated levels of gene transcripts originating from the brain, compared to plasma. Moreover, the CSF profile of AD subjects appears to recapitulate the reported biological changes associated with AD and may provide insights into other disruptions of brain homeostasis. Collectively, our data suggests that CSF cf-mRNA profiling serves as a valuable surrogate for evaluating the molecular dysregulation occurring within the brain.

We were surprised in our previous study by the relatively large contribution of brain associated cf-mRNA transcripts in the circulation (15). Our initial thoughts were that the central role of the brain in organismal biology may explain the apparent overrepresentation in the plasma cf-mRNA compartment. Upon further consideration, we are struck by the various biological exchanges across the three ‘barriers’, or perhaps more accurately ‘interfaces’, of communication between the brain and the periphery, namely the Blood Brain, Outer Brain (arachnoid) and Blood CSF. Transport across these ‘barriers’ occurs by passive diffusional as well as active transport mechanisms, and disruptions of these barriers occur in the pathophysiology of progressive disease. The blood-brain barrier and blood-CSF barrier serve as the largest interfaces connecting blood and brain extracellular fluids (23). Furthermore, considering that these interfaces, particularly the blood-brain barrier, are recognized for effectively clearing a substantial portion of AD-related forms of Aβ (59), disruption or dysregulation of these barriers is closely associated with AD, and other neurodegenerative diseases (60, 61). Functionally, these barriers offer distinct pathways for transporting brain-derived molecules into the circulation (62). In addition, given that a substantial portion of brain-derived molecules enter circulation, it is plausible that a significant amount of brain-derived transcripts could be from CSF rather than being directly released through the blood-brain barrier. Considering the potential of blood as a source for developing non-invasive biomarkers, further research is required to understand the underlying mechanism in which brain-derived transcripts are released into the circulation.

Two of the limitations of our study are the small sample size and lack of an independent validation set. Also, late-stage disease testing may not capture a ‘natural history ledger’ for the pathological continuum of underlying disease mechanisms. For future studies, it will be crucial to conduct research with CSF samples collected at various stages of disease, with sufficient sample size, to gain a more comprehensive understanding of the AD continuum. In conclusion, we conducted comprehensive cf-mRNA profiling in CSF samples obtained from subjects with AD and NCI and identified dysregulated AD genes consistent with the present understanding of pathogenesis. We further developed cf-mRNA CSF disease classifiers with the potential to inform future blood-based AD diagnostic tools. Our results highlight the possibility of CSF and plasma cf-mRNA profiling as an alternative to the current standard of care and invasive procedures, offering a means to investigate novel molecular dysregulation within the brain.

## Data Availability

All data produced in the present study are available upon reasonable request to the authors

## ACKNOWLEDGEMENT

We thank Drs. Michael Nerenberg and Arkaitz Ibarra for their contributions to the initial study design. We also thank the technical assistance provided by Amy Karns and Alex Acosta.

## AUTHOR CONTRIBUTIONS

R.E.D.S., J.J.S. and S.T. conceived and designed manuscript; R.E.D.S., S.K. and S.T. prepared figures; R.E.D.S., J.J.S. and S.T. drafted manuscript; R.E.D.S., S.K., S.R.Q, J.J.S. and S.T. edited and revised manuscript.

