## Supplementary Material for "Transcriptome profiling of cerebrospinal fluid in Alzheimer’s Disease reveals molecular dysregulations associated with disease"

#### Supplementary Figure 1

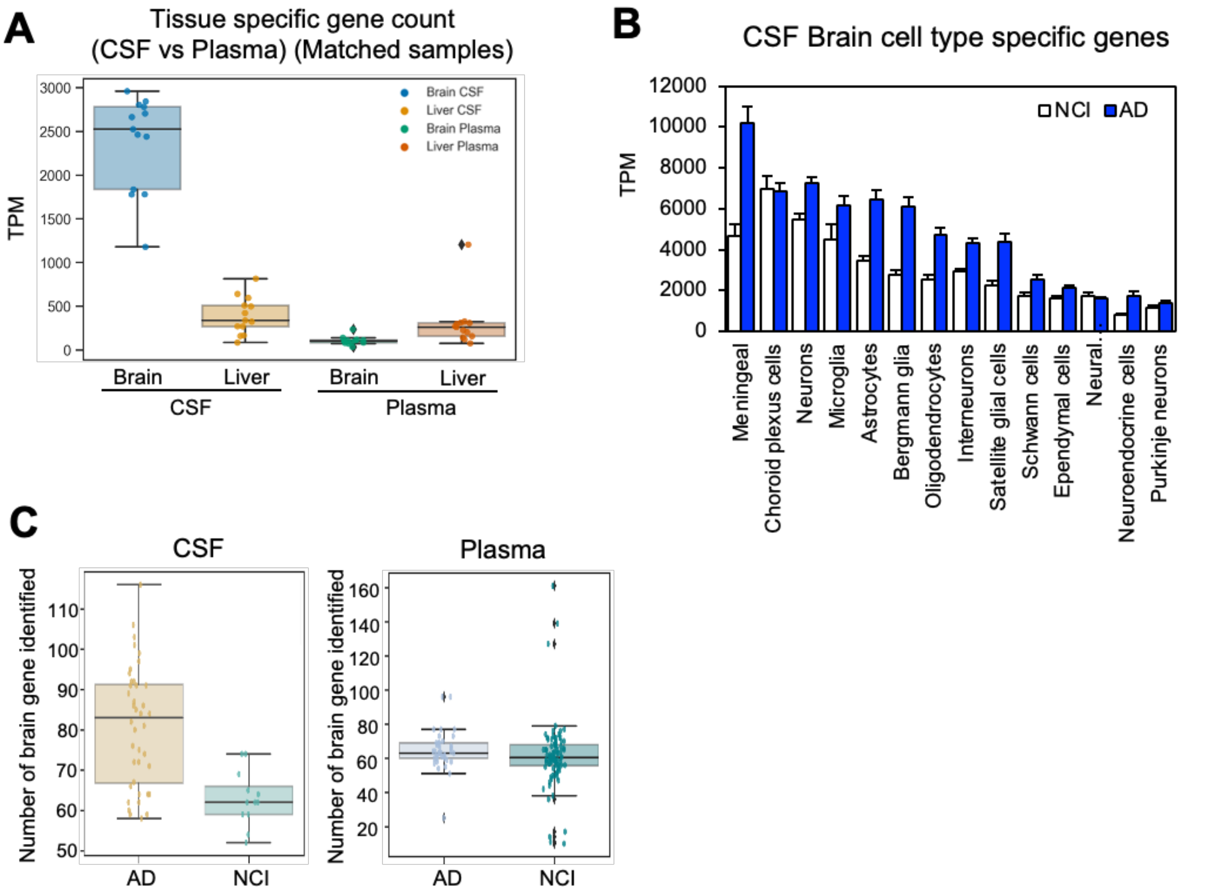


**Supplementary Figure 1:** Brain cell and tissue type specific genes in CSF and plasma. A) Average brain and liver specific transcripts for CSF and plasma (Matched samples only). B) Estimation of brain cell types in CSF between AD and NCI. C) Number of brain genes identified in CSF (left) and plasma (right) for AD and NCI.

### Supplementary Figure 2

**A**

| Most Differentially Expressed Genes | | | | | |
|---|---|---|---|---|---|
| Upregulation | FC | P-value | Downregulation | FC | P-value |
| CKB | 1.58 | 0.00004 | MCTS1 | -2.64 | 0.00004 |
| NBL1 | 2.20 | 0.00006 | MBD4 | -1.99 | 0.00007 |
| IGF2 | 1.24 | 0.0002 | PPIA | -0.88 | 0.00009 |
| CDH7 | 5.04 | 0.0003 | SMC3 | -1.48 | 0.0001 |
| TMEM63C | 5.65 | 0.0003 | SRP72 | -1.59 | 0.0001 |
| SLC2A1 | 3.11 | 0.0005 | MED4 | -1.59 | 0.0001 |
| HMG20B | 1.38 | 0.0005 | GLRX | -1.73 | 0.0002 |
| RNH1 | 1.51 | 0.0005 | RPL36A | -1.02 | 0.0002 |
| ARHGEF15 | 4.34 | 0.0005 | RPL30 | -0.91 | 0.0003 |
| ZNF665 | 2.06 | 0.0005 | BCLAF1 | -1.45 | 0.0003 |

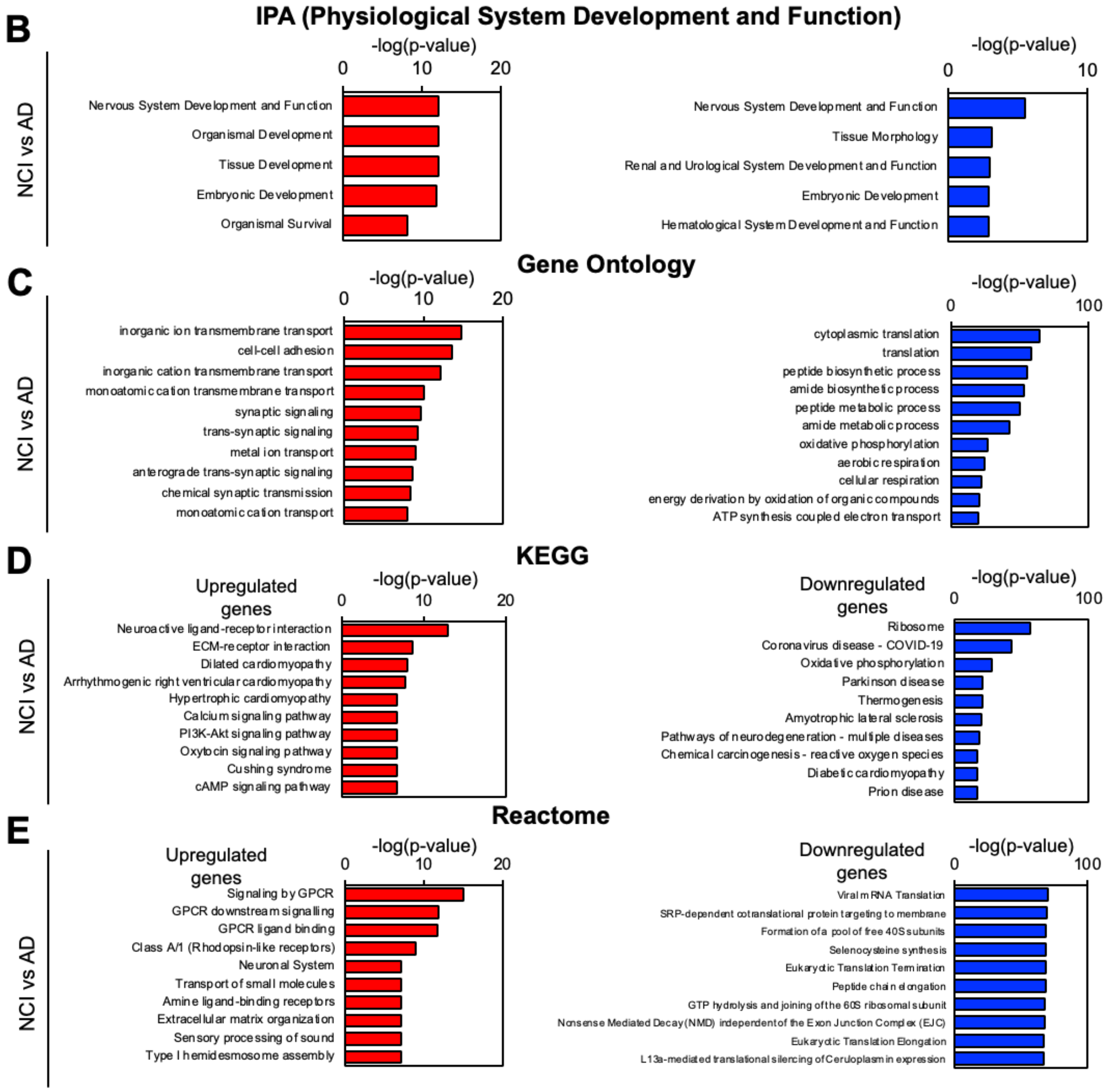

**Supplementary Figure 2:** Differentially expressed CSF cf-mRNA genes between AD and NCI. A) The list of top 10 most upregulated genes (left) and downregulated genes (right) ranked by p-value. B) Top IPA Physiological System Development and Function pathways for genes that are dysregulated in AD, upregulated and downregulated genes are used as input. C) Top Gene Ontology pathways for genes that are dysregulated in AD, upregulated and downregulated genes are used as input. D) Top KEGG pathways for genes that are dysregulated in AD, upregulated and downregulated genes are used as input. E) Top Reactome pathways for genes that are dysregulated in AD, upregulated and downregulated genes are used as input.

#### Supplementary Figure 3

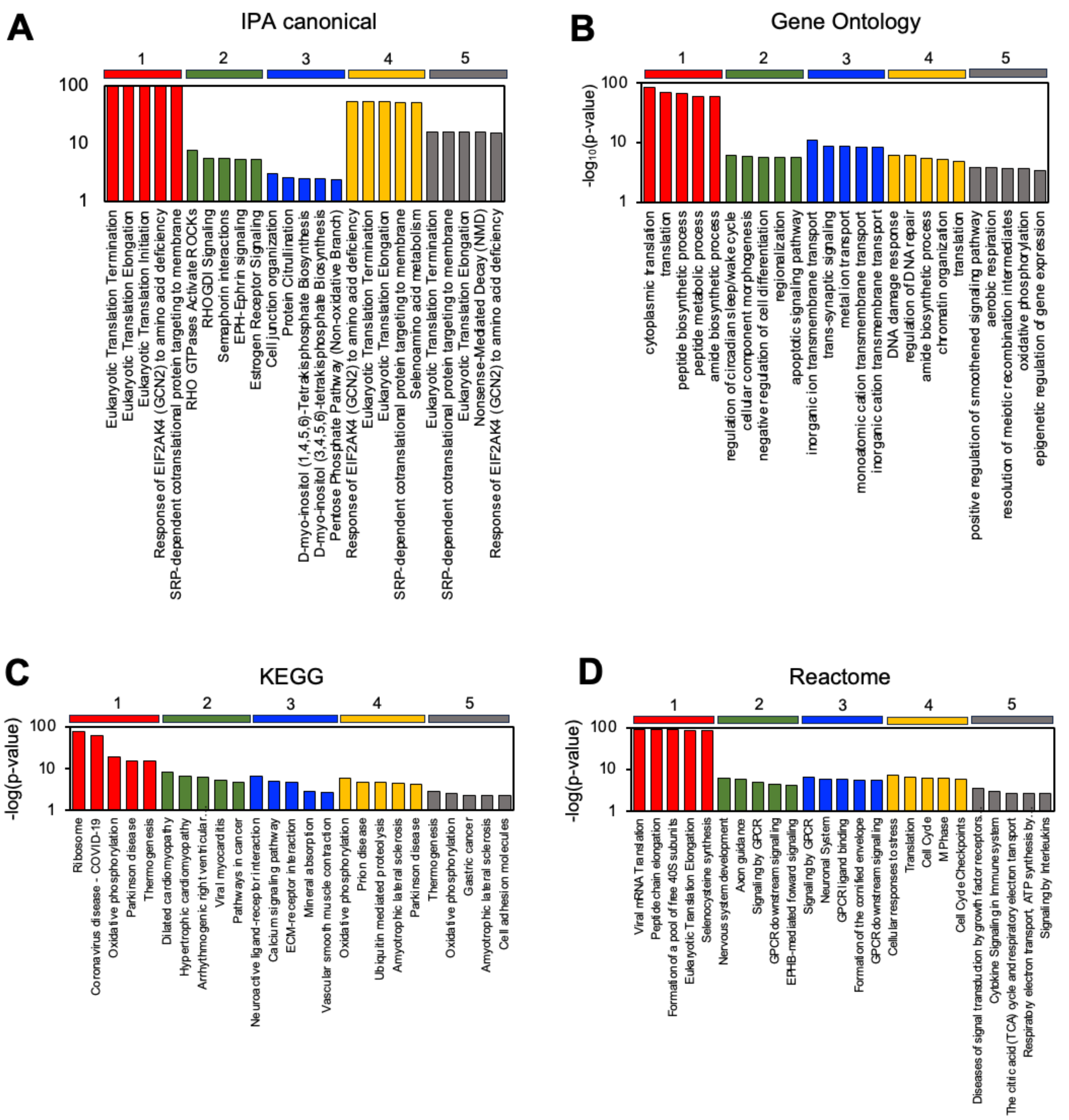


**Supplementary Figure 3:** NMF analysis for AD differentially expressed genes. A) Top five IPA canonical pathways for each cluster generated from NMF analysis (AD vs NCI). B) Top five Gene Ontology pathways for each cluster generated from NMF analysis (AD vs NCI). C) Top five Reactome pathways for each cluster generated from NMF analysis (AD vs NCI). D) Top five KEGG pathways for each cluster generated from NMF analysis (AD vs NCI).

### Supplementary Figure 4

**A**

| Most Differentially Expressed Genes | | | | | |
|---|---|---|---|---|---|
| Upregulation | FC | P-value | Downregulation | FC | P-value |
| FBXO9 | 1.50 | 7.32E-05 | CNR1 | -4.45 | 0.001 |
| SEZ6L2 | 2.81 | 0.0002 | GPD2 | -3.06 | 0.002 |
| CCT6B | 1.81 | 0.0003 | GOLGA3 | -1.91 | 0.003 |
| IQCF6 | Not ex | 0.0004 | C1orf21 | -2.84 | 0.003 |
| AIRN | Not ex | 0.0004 | PSPC1 | -3.98 | 0.004 |
| TMTC4 | 2.32 | 0.0008 | GDF11 | -1.87 | 0.005 |
| PLEKHG3 | 2.12 | 0.0008 | SIKE1 | -2.39 | 0.005 |
| GNG10 | 1.43 | 0.0009 | PAN2 | -2.43 | 0.005 |
| ANKRD36C | 3.63 | 0.001 | FYN | -2.39 | 0.006 |
| ATP2B2 | 2.11 | 0.001 | MYADM | -4.34 | 0.006 |

**B**

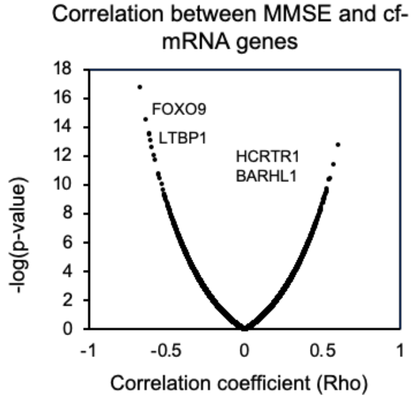


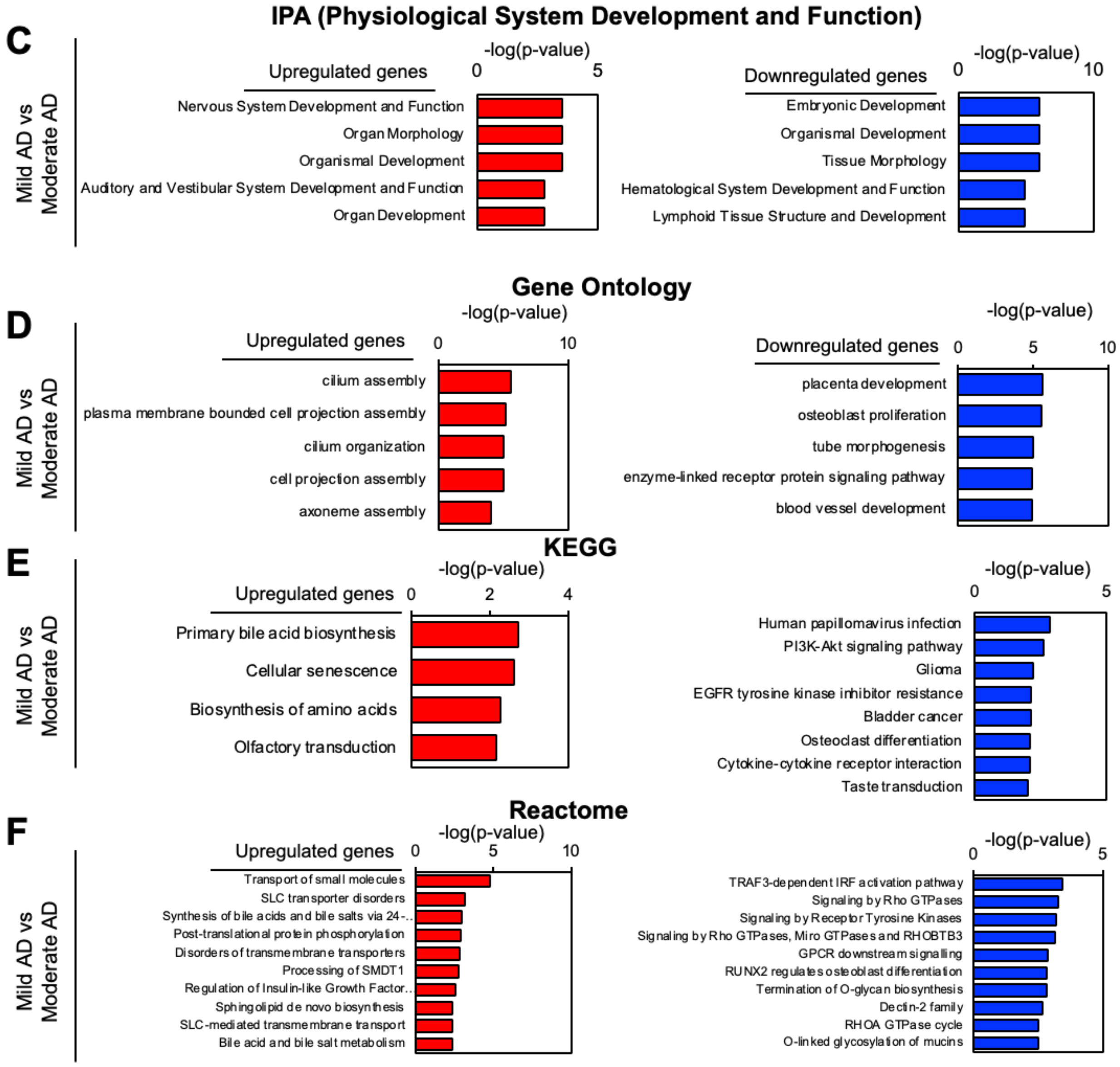

**Supplementary Figure 4:** Differentially expressed CSF cf-mRNA genes between mild AD and moderate AD. A) The list of top 10 most upregulated genes (left) and downregulated genes (right) ranked by p-value (upregulated means genes that are overexpressed in moderate AD compared to mild AD). B) Correlation between MMSE and cf-mRNA genes. The graph plots p-value against correlation coefficient. C) Top IPA Physiolological System Development and Function pathways for genes that are dysregulated in moderate AD compared to mild AD, upregulated and downregulated genes are used as input. D) Top Gene Ontology pathways for genes that are dysregulated in moderate AD compared to mild AD, upregulated and downregulated genes are used as input. E) Top KEGG pathways for genes that are dysregulated in moderate AD compared to mild AD, upregulated and downregulated genes are used as input. F) Top Reactome pathways for genes that are dysregulated in moderate AD compared to mild AD, upregulated and downregulated genes are used as input.

#### Supplementary Figure 5

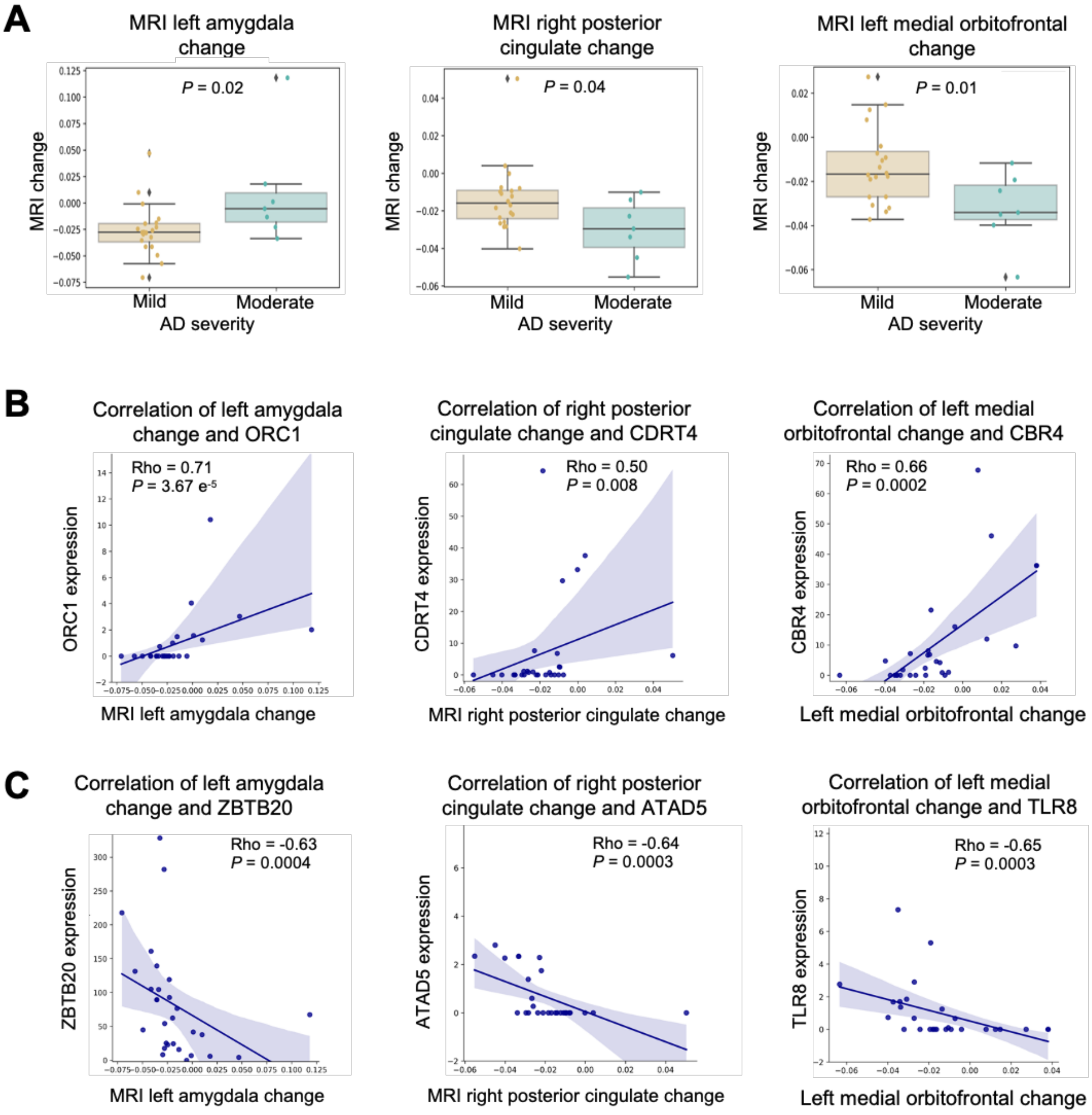


Supplementary Figure 5: MRI measurements and cf-mRNA changes. A) MRI measurements between mild and moderate AD (left: left amygdala, middle: posterior cingulate, right: right medial orbitofrontal). B) Genes that positively correlated most with MRI measurements (left: left amygdala, middle: posterior cingulate, right: right medial orbitofrontal). C) Genes that negatively correlated most with MRI measurements (left: left amygdala, middle: posterior cingulate, right: right medial orbitofrontal).

#### Supplementary Figure 6

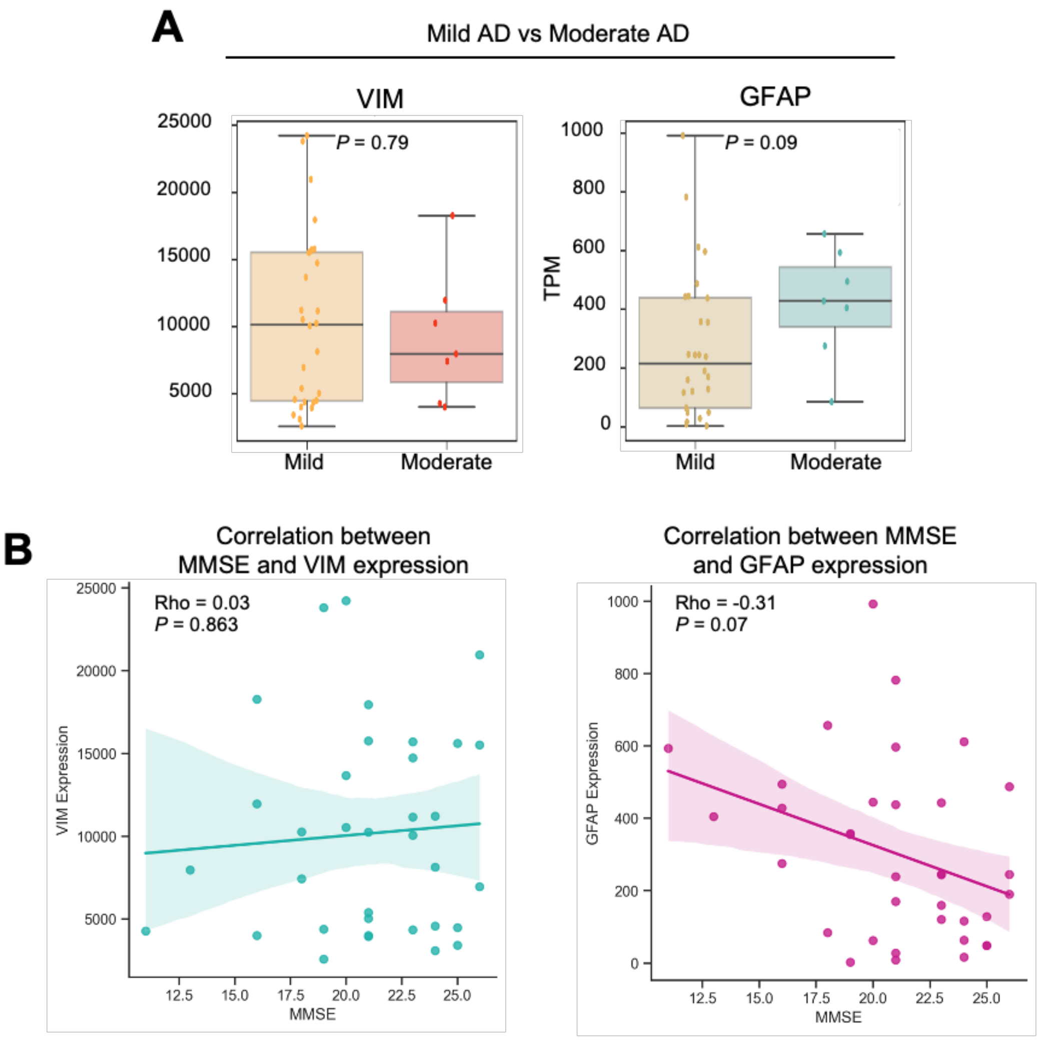


Supplementary Figure 6: Astrocyte Reactivity biomarkers in CSF. A) The expression levels of astrocyte activity markers in CSF (moderate AD vs mild AD). B) Correlation analysis between astrocyte markers in CSF against MMSE score.

#### Supplementary Figure 7

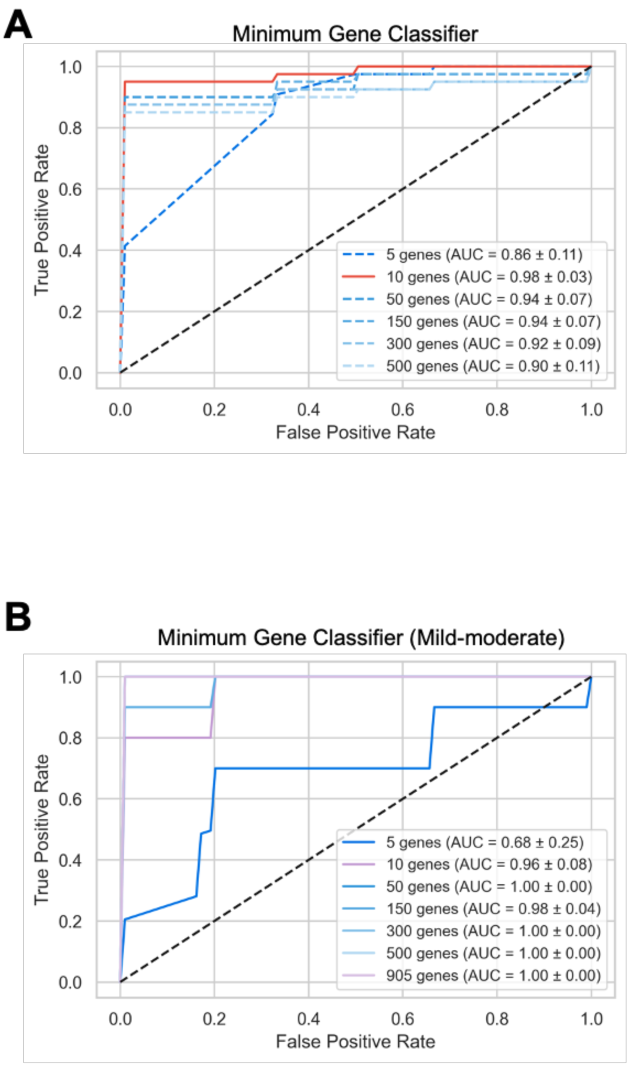


**C**

| Ranking | Gene |
|---|---|
| 1 | ENSG00000104964.14 |
| 2 | ENSG00000136521.12 |
| 3 | ENSG00000103145.10 |
| 4 | ENSG00000092330.15 |
| 5 | ENSG00000102879.15 |
| 6 | ENSG00000126822.16 |
| 7 | ENSG00000168000.14 |
| 8 | ENSG00000180573.9 |
| 9 | ENSG00000130313.6 |
| 10 | ENSG00000198300.12 |
| 11 | ENSG00000182196.13 |
| 12 | ENSG00000169223.14 |
| 13 | ENSG00000108821.13 |
| 14 | ENSG00000197183.12 |
| 15 | ENSG00000143252.14 |
| 16 | ENSG00000126934.13 |
| 17 | ENSG00000159176.13 |
| 18 | ENSG00000171148.13 |
| 19 | ENSG00000159212.12 |
| 20 | ENSG00000163875.15 |
| 21 | ENSG00000235718.8 |
| 22 | ENSG00000013364.18 |
| 23 | ENSG00000108840.15 |
| 24 | ENSG00000010810.17 |
| 25 | ENSG00000104805.15 |
| 26 | ENSG00000185507.19 |
| 27 | ENSG00000184922.13 |
| 28 | ENSG00000185359.12 |
| 29 | ENSG00000112146.16 |
| 30 | ENSG00000169554.16 |
| 31 | ENSG00000079313.11 |
| 32 | ENSG00000138083.4 |
| 33 | ENSG00000149798.4 |
| 34 | ENSG00000140943.16 |
| 35 | ENSG00000171223.5 |
| 36 | ENSG00000197345.12 |
| 37 | ENSG00000116786.11 |
| 38 | ENSG00000125734.13 |
| 39 | ENSG00000005022.5 |
| 40 | ENSG00000184060.10 |
| 41 | ENSG00000169228.13 |
| 42 | ENSG00000130600.15 |
| 43 | ENSG00000100568.10 |
| 44 | ENSG00000119559.15 |
| 45 | ENSG00000198771.10 |
| 46 | ENSG00000113719.15 |
| 47 | ENSG00000198837.9 |
| 48 | ENSG00000038382.17 |
| 49 | ENSG00000109171.14 |
| 50 | ENSG00000002822.15 |

Supplementary Figure 7: Establishing gene-classifiers for AD diagnosis. A) AD diagnostic gene classifiers using different number of genes as input. B) Diagnostic gene classifier to distinguish mild AD from moderate AD using different number of genes as input. C) The list of 50 genes used to for mild-moderate AD diagnostic classifier.

#### SUPPLEMENTARY TABLES

**Supplementary Table 1:** Subject characteristics table

| **Disease** | | AD | NCI |
|---|---|---|---|
| n | | 40 | 12 |
| Age | Average (± SEM) | 74.4 ± 1.2 | 68.3 ± 1.6 |
| Sex | Female (%) | 23 (57.5%) | 7 (58.3%) |
| | Male (%) | 17 (42.5%) | 5 (41.7%) |
| Cognitive impairment | Average MMSE score (± SEM) | 20 .9 ± 0.6 | NA |

**Supplementary Table 2:** The list of brain and liver specific genes

**Brain**

| | | | | | |
|---|---|---|---|---|---|
| AC000067.1 | CAMKV | GLRA2 | LL0XNC01-16G2.1 | RIT2 | RP5-1180C18.1 |
| AC004603.4 | CBLN1 | GNG13 | LRTM2 | RP1-223B1.1 | RP5-860P4.2 |
| AC005392.13 | CCK | GPR101 | MAP6D1 | RP1-257I9.2 | RPRML |
| AC006041.1 | CDH22 | GPR62 | MCHR2 | RP1-269M15.3 | SCN2A |
| AC007106.1 | CENPVP1 | GPR88 | MEPE | RP11-107I14.5 | SLC12A5 |
| AC008060.7 | CHN1 | GRIA4 | MGAT5B | RP11-142A5.1 | SLC17A7 |
| AC008060.8 | CNPY1 | GRIN1 | MIR381HG | RP11-148B3.1 | SLC1A2 |
| AC009110.1 | CNTNAP4 | GRM1 | MLC1 | RP11-149P24.1 | SLC24A2 |
| AC009227.3 | CRTAM | GRM3 | MOBP | RP11-157E21.1 | SLC39A12 |
| AC011897.2 | CSPG5 | GRM4 | MOG | RP11-167H9.3 | SLC6A3 |
| AC011995.1 | CTA-929C8.8 | GS1-519E5.1 | NCAN | RP11-173E2.2 | SLCO1A2 |
| AC062021.1 | CTA-992D9.7 | GSX1 | NCDN | RP11-231C18.1 | SNAP25 |
| AC068057.1 | CTB-1I21.1 | HAPLN2 | NEUROD1 | RP11-231C18.2 | SNCB |
| AC068490.1 | CTC-472C24.1 | HCRT | NEUROD2 | RP11-231N3.1 | SRRM4 |
| AC068535.2 | CTD-2009A10.1 | HPCA | NEUROD6 | RP11-232M24.1 | STMN2 |
| AC140542.2 | CTD-2228A4.1 | HRH3 | NKX6-2 | RP11-238K6.2 | STMN4 |
| AC140912.1 | CTD-2316B1.1 | HTR1A | NMS | RP11-25O3.1 | STX1B |
| AF240627.2 | CTD-2316B1.2 | HTR2C | NPVF | RP11-268E23.2 | SYNDIG1L |
| AJ003147.8 | CTD-2533K21.4 | HTR5A | NR2E1 | RP11-298J23.8 | SYNPR |
| AL050303.7 | CTD-2555A7.2 | IFNA8 | NTSR2 | RP11-2L8.2 | SYT1 |
| ANO3 | DIRAS2 | KCNA1 | OLIG2 | RP11-331K15.1 | TBR1 |
| AP000797.3 | DRD1 | KCNC1 | OPALIN | RP11-33M22.2 | TFAP2D |
| ATP6V1G2 | ELAVL3 | KCND2 | OPCML | RP11-354K1.1 | TH |
| AVP | EN2 | KCNJ9 | OR2M4 | RP11-375B1.2 | TLX3 |
| BARHL1 | ERMN | KCNK4 | OR5J2 | RP11-399H11.3 | TMEM151B |
| BCAN | FAM131B | KCNK9 | OTP | RP11-421P23.2 | TMEM178A |
| BRINP1 | FAM163B | KCNN1 | OTX2 | RP11-429O1.1 | TMEM235 |
| C11orf87 | FBXL16 | KCNV1 | OXT | RP11-430H10.3 | TMEM88B |
| C18orf42 | FGF3 | KIF5A | PACSIN1 | RP11-430H10.4 | TRIM67 |
| C1orf61 | FSTL5 | KLHL1 | PAQR6 | RP11-449H15.2 | TTC9B |
| C1QL2 | GABBR2 | LAMP5 | PAX6 | RP11-458K10.1 | UNC13C |
| C1QL3 | GABRA1 | LHX2 | PDYN | RP11-491F9.5 | UNCX |
| C2orf80 | GABRA3 | LINC00290 | PLP1 | RP11-491F9.6 | VSNL1 |
| C9orf62 | GABRA6 | LINC00320 | POU3F4 | RP11-491F9.8 | VSTM2A |
| CA10 | GABRB2 | LINC00387 | PPFIA4 | RP11-587P21.3 | ZIC1 |
| CABP1 | GABRD | LINC00460 | PSD2 | RP11-805F19.1 | ZIC2 |
| CACNA1A | GABRG1 | LINC00599 | PTPN5 | RP11-90J7.2 | ZIC3 |
| CACNG3 | GAD2 | LINC01007 | PTPRT | RP11-953B20.1 | ZIC4 |
| CACNG8 | GAP43 | LINC01034 | RESP18 | RP11-981G7.6 | ZP2 |
| CAMK2A | GFAP | LINGO1 | RIMS1 | RP5-1177M21.1 | |

**Liver**

| | | | | | |
|---|---|---|---|---|---|
| ABCG5 | APOC2 | CPB2 | FGG | MBL2 | SERPINA7 |
| AC003988.1 | APOC4 | CRP | GC | OR10J5 | SERPINC1 |
| AC006037.2 | APOF | CTD-2526M8.2 | GLYATL3 | ORM1 | SERPIND1 |
| AC068535.3 | APOH | CTD-2582M21.1 | HAO1 | ORM2 | SLC10A1 |
| ACOT12 | ASGR1 | CTD-3162L10.4 | HP | PLG | SLC13A5 |
| ADH1A | ASGR2 | CYP2A6 | HPX | PON1 | SLC17A2 |
| ADH4 | BAAT | CYP2C8 | HRG | PROC | SLC25A47 |
| AFM | C8A | CYP2E1 | HULC | RDH16 | SLCO1B1 |
| AGXT | C8B | CYP8B1 | IGFBP1 | RP11-101E14.3 | SLCO1B3 |
| AHSG | C8G | F13B | INHBC | RP11-1151B14.2 | TAT |
| AKR1C4 | C9 | F2 | INHBE | RP11-622A1.2 | TDO2 |
| ALB | CFHR1 | F7 | ITIH1 | RP11-685F15.1 | U91324.1 |
| AMBP | CFHR2 | F9 | ITIH2 | RP11-753B14.1 | UGT1A4 |
| ANGPTL3 | CFHR3 | FAM99A | ITIH4 | SAA4 | UGT2B10 |
| APCS | CFHR4 | FGA | LBP | SERPINA10 | UGT2B4 |
| APOA2 | CFHR5 | FGB | LPA | SERPINA11 | UROC1 |
| APOA5 | COLEC10 | FGF21 | MAT1A | SERPINA6 | |
